# Pigmented Paraganglioid Carcinoid Tumors of the Lung: Spatial Transcriptomics Reveals Shared and Distinct Features with Typical Carcinoid Tumors and Extra-Adrenal Paragangliomas

**DOI:** 10.64898/2025.11.29.25341268

**Authors:** Hisham F. Bahmad, Alfredo Perez-Tagle-Tejeda, Brenda M. Cisneros-Gonzalez, Rodrigo Santoscoy-Valencia, Jessica Alvarez-Lesmes, Katherine Drews-Elger, Laurence M. Briski, Manuel Lora-Gonzalez, Andre Pinto, Andrew E. Rosenberg, Roberto Ruiz-Cordero

**Affiliations:** Jackson Memorial Hospital, Miami, FL 33136, USA; Department of Internal Medicine, Pembroke Pines, FL 33028, USA; Pediatrics, Memorial Healthcare System, Pembroke Pines, FL 33028, USA; Baptist Health System of South Florida, Miami, FL 33176, USA; Department of Pathology and Laboratory Medicine, University of Miami Miller School of Medicine, Miami, Florida 33136, USA; Sylvester Cancer Center, University of Miami Miller School of Medicine, Miami, Florida 33136, USA

**Keywords:** pigmented paraganglioid carcinoid tumor, typical carcinoid tumor, paraganglioma, pulmonary neuroendocrine tumor, melanocytic differentiation, digital spatial profiling, transcriptomics, immunohistochemistry

## Abstract

Pigmented paraganglioid carcinoid tumors (PPCT) of the lung are a rare, underrecognized, and poorly characterized morphologic variant of pulmonary neuroendocrine tumors (NETs). While these tumors are usually diagnosed as typical carcinoid (TC) tumors, PPCT may represent a diagnostic challenge due to the histologic resemblance with extra-adrenal paraganglioma (PG). In this study, we aimed to comprehensively characterize the histomorphologic, immunophenotypic, and transcriptomic profiles of PPCT in comparison to TC and PG using spatially resolved transcriptomic analysis. Using a tissue microarray (TMA) composed of 38 tumors, including 20 TC, 16 PG, and 2 PPCT, we performed immunohistochemical (IHC) and digital spatial transcriptomic (GeoMx® DSP) profiling. The TMA included two punches and two regions of interest (ROIs) per case. Cellular transcriptomes were selected based on epithelial (PanCK+), sustentacular (S100+), and immune (CD45+) compartments. By IHC, PPCT retained neuroendocrine markers (synaptophysin, INSM1, chromogranin A) but showed decreased or absent pancytokeratin cocktail expression and increased number of sustentacular cells highlighted by strong expression of S100 and SOX-10, similar to PG. Expression of AE1/AE3 and CK8/18 confirmed their epithelial origin and helped distinguish them from PG. The transcriptome of PPCT clustered with that of TC but displayed distinct expression patterns in a small subset of genes. Although the sustentacular and immune compartments showed limited divergence, the epithelial compartment showed differentially expressed genes in PPCT including *FABP5*, *MLPH*, *GPNMB*, and *SOX1*, which indicate upregulation of melanocytic and neural crest markers. Gene set enrichment analysis (GSEA) revealed significant upregulation of pathways related to inflammation (e.g., *TLR4*-*TRAF6*-*TAK1*), *PTEN* trafficking, and inositol phosphate metabolism. PPCT show increased melanocytic pathway expression, which may explain the morphologic resemblance to PG.

## 1. Introduction

Pulmonary carcinoid tumors of the lung account for ≤ 2% of all primary lung malignancies ^1–4^. These tumors are categorized by the World Health Organization (WHO) classification into two major subtypes: typical carcinoids (TC) and atypical carcinoids (ACs), distinguished primarily by mitotic activity and the presence or absence of necrosis ^4^. Whereas TC are defined by < 2 mitoses per 2 mm^2^ and the absence of necrosis, ACs are characterized by 2 to 10 mitoses per 2 mm^2^ and/or the presence of necrosis (usually punctate or focal). TC are considered low-grade carcinomas that often present in younger patients, and while they exhibit an indolent clinical behavior, they harbor metastatic potential. ACs, on the other hand, represent intermediate-grade tumors with more aggressive clinical behavior, higher propensity for metastasis, and less favorable outcomes ^4–6^. A limited number of case reports are available in the literature that document a rare variant of this tumor that has been referred to as paraganglioid carcinoid tumors ^7–10^, pigmented paraganglioid carcinoid ^11–14^ or peripheral melanotic paraganglioid carcinoid ^14–19^.

Pigmented paraganglioid carcinoid tumors (PPCT) represent a morphologic variant histologically defined by a zellballen-like nested architecture and the presence of increased S100+ sustentacular cells, resembling paragangliomas (PG) ^7–13,15–19^. Reports have noted that the S100+ cells in PG and some pulmonary neuroendocrine tumors (NETs) represent Schwann-cell-like cells, which provide structural and trophic support to the tumor microenvironment ^20,21^. They also play a role in maintaining the architectural integrity of zellballen structures and may influence tumor differentiation and immune modulation. The terms “pigmented carcinoid”, “melanocytic carcinoid”, and “melanin-containing carcinoid” are often used when tumor cells contain abundant intracytoplasmic pigment granules that are coarsely granular and brownish in appearance, typically representing neuromelanin ^11–19^. Historically, Capella *et al*. (1979) described a subset of 11 bronchial carcinoids exhibiting paraganglioid morphology with ultrastructural similarities to pulmonary P cells ^7^. In another study by Reil *et al*. (1998), 10 tumors with histologic and immunophenotypic resemblance to paragangliomas were classified into typical pulmonary paraganglioid carcinoids and atypical pulmonary paraganglioid carcinoids categories following Arrigoni criteria ^8,9,17,22^.

PG are non-epithelial, neuroendocrine tumors arising from paraganglia associated with the autonomic nervous system and are most frequently found in the adrenal medulla or head and neck region ^23–25^. Extra-adrenal PG of the thorax, including those of the lung, are exceedingly rare but must be considered in the differential diagnosis when encountering pigmented neuroendocrine neoplasms with a zellballen architecture ^23,26–31^. The key challenge in distinguishing PPCT from extra-adrenal PG in the lung lies in their overlapping histomorphological and immunohistochemical features ^32,33^. Both tumors express neuroendocrine markers such as synaptophysin and chromogranin A, and both may exhibit S100 positivity in a sustentacular pattern ^4,32–34^. Additionally, both may express SOX-10 by immunohistochemistry (IHC) ^4,35^, a transcription factor involved in neural crest development, which is widely expressed in PG and melanocytic lesions ^36,37^. However, the epithelial origin of carcinoid tumors often results in low-molecular-weight cytokeratin (CK) positivity, such as CK8/18 and AE1/AE3, while PG are typically negative for these markers ^38^. The origin of pulmonary carcinoid tumors is believed to arise from neuroendocrine Kulchitsky cells located in the bronchial epithelium ^39,40^. PPCT, however, possess additional features reminiscent of neural crest-derived paragangliomas, including their cellular composition (neuroendocrine cells and sustentacular cells) and zellballen-like architectural patterns.

At the molecular level, no definitive driver mutations have yet been identified in PPCT that distinguish them from conventional carcinoids or extra-adrenal PG. The IHC expression of BCL-2 protein has been noted in all pulmonary neuroendocrine tumors regardless of subtype ^9^, indicating it is not a discriminative biomarker. PPCT share indolent behavior with conventional TC. For instance, in the series by Resl *et al*. (1998), eight tumors were available for follow-up showing no recurrence in a 1 to 12-year period post-resection^9^. Despite the phenotypic overlap between PPCT and both conventional carcinoid tumors and PG, the true incidence, pathogenesis, and molecular landscape of PPCT remain inadequately explored. Recent spatial transcriptomic advances offer opportunities to delineate the molecular landscape of these tumors, particularly in identifying melanocytic lineage genes and Schwannian support cells contributing to their phenotype. To address these knowledge gaps, our present study systematically characterizes the histomorphologic, immunophenotypic, and transcriptomic profiles of PPCT, and compares them to pulmonary carcinoid tumors and extra-adrenal PG.

## 2. Materials and Methods

### 2.1. Study Design and Sample Selection

This is a retrospective study conducted on surgical resection specimens including TC of the lung, extra-adrenal PG, and PPCT of the lung. Archival pathology specimens were retrieved from the pathology files at the University of Miami between January 2021 and August 2022. Inclusion criteria included availability of formalin-fixed paraffin-embedded (FFPE) tissue, complete clinical data, and adequate diagnostic tissue for tissue microarray (TMA) construction. Exclusion criteria included inadequate tissue for immunohistochemical and transcriptomic analyses or incomplete patient records. Clinical and demographic data were extracted for all patients from the medical charts (**Supplementary Table S1**). Patient information included sex, age at diagnosis, tumor site, smoking status, race/ethnicity, tumor size, and stage (pT, pN), when applicable.

### 2.2. Ethical Considerations

This study was approved by the Institutional Review Board (IRB) of the University of Miami Miller School of Medicine (IRB# 20180047). All patient data were de-identified to maintain confidentiality, and the study was performed in accordance with the Declaration of Helsinki and institutional ethical standards.

### 2.3. Tissue Microarray (TMA) Construction

Representative tumor areas were selected by two pathologists (RRC and LMB) and punched in duplicate (1-mm cores) to construct a TMA (**Supplementary Fig. S1**). The resulting TMA block included two cores per tumor to ensure reproducibility and account for intratumoral heterogeneity. Unstained 4-µm sections were prepared from the TMA block for subsequent IHC and transcriptomic analysis.

### 2.4. Immunohistochemistry (IHC) and Evaluation of Immunohistochemical Expression

IHC was performed on the Ventana Benchmark ULTRA automated staining system (Ventana Medical Systems, Tucson, AZ, USA) using the following antibodies: pancytokeratin (PanCK) cocktail, AE1/AE3, CK8/18, synaptophysin, chromogranin A, INSM1, TTF-1, GATA-3, S100, SOX-10, and Ki-67. Antibody dilutions, retrieval conditions, and detection systems followed the manufacturer’s standard protocols. Staining patterns were independently assessed by two board-certified pathologists (RRC and LMB). The extent of staining was categorized as diffuse (>50% tumor cells), intermediate (11–50%), focal (1–10%), or negative (0%) for PanCK, AE1/AE3, CK8/18, synaptophysin, chromogranin A, INSM1, TTF-1, and GATA-3. S100 and SOX-10 were evaluated for presence or absence of sustentacular staining. Ki-67 labeling indices were manually estimated as percentages of positively stained tumor nuclei.

### 2.5. Digital Spatial Profiling (GeoMx® DSP)

We employed the NanoString GeoMx® Digital Spatial Profiling (DSP) to assess the spatial transcriptomic landscapes of PPCT and compare the results to those of TC and PG (**Figure 1**). The GeoMx® DSP platform (NanoString Technologies, Inc.) allows for comprehensive profiling of mRNA targets in FFPE tissue sections, providing insights into tumor heterogeneity, immune microenvironments, and differentially expressed genes and signaling pathways. FFPE slides were submitted per NanoString guidelines, and regions of interest (ROIs) were selected based on hematoxylin and eosin (H&E) and immunofluorescence staining patterns.

**Figure 1.**
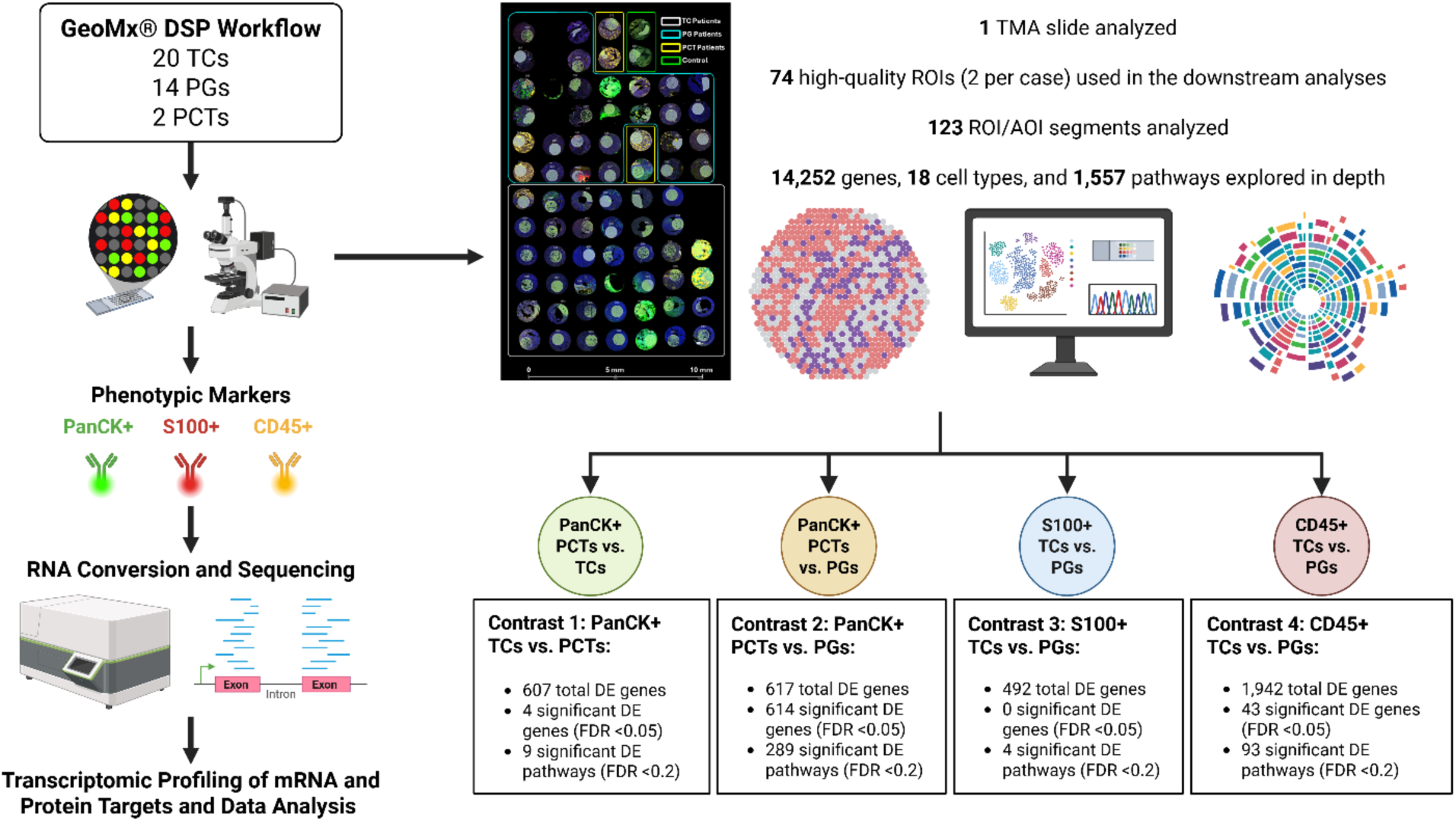
Schematic flow chart of the GeoMx® DSP workflow applied to TC, PG, and PPCT. The schematic was created in BioRender. Bahmad, H. (2025) https://BioRender.com/193fd0b. Abbreviations: AOI: area of illumination; DE: differentially expressed; DSP: Digital Spatial Profiler; FDR: false discovery rate; PanCK: pancytokeratin cocktail; PCT: pigmented or paraganglioid carcinoid tumor; PG: paraganglioma; ROI: region of interest; TC: typical carcinoid; TMA: tissue microarray.

Quality control (QC) was performed to ensure integrity and reliability of the DSP data, metrics include sequencing saturation (>50%), aligned reads, and appropriate probe detection thresholds. Two PG cases (Patients 8 and 15) failed quality control (QC) and were excluded. Thus, the final cohort for spatial transcriptomic profiling included 20 TC, 2 PPCT, and 14 PG, along with one normal heart case included for orientation. GeoMx® DSP was conducted using the Whole Transcriptome Atlas (WTA) panel targeting over 18,000 protein-coding genes. ROIs were selected based on tumor-rich areas, excluding necrotic or artifactual regions, resulting in a total of 74 high-quality ROIs (two per case) used for downstream analyses.

Data were acquired and analyzed using GeoMx® DSP software and nSolver analysis suite. The remaining dataset showed robust detection with an average of >80% of targets passing filtering thresholds across ROIs.

In summary, 128 Area of Illumination (AOI) segments were profiled and 123 passed QC. Three AOIs were flagged for low area, and 2 were flagged for low gene detection rate. A total of 14,252 genes were detected in at least 30% of segments and were analyzed further. Each ROI/AOI segment has several unique negative control probes that in aggregate can be used to estimate background (**Supplementary Fig. S1**). Outlier negative control probes were removed from the data – either entirely (global) or from specific segments (local) – to refine the estimation of background and downstream gene detection. Data underwent third quartile (Q3) normalization. The purpose of normalization is to adjust for technical variables, such as ROI/AOI surface area and tissue quality, and enable meaningful biological and statistical discoveries. Two common methods for normalization of GeoMx® WTA data are i) Q3 or ii) background. Both methods estimate a normalization factor per ROI/AOI segment to bring the segment data distributions together. Q3 is typically the preferred approach.

Spatial deconvolution analysis was employed to estimate the relative abundance of cell types for each ROI/AOI segment. The algorithm utilizes a pre-specified cell profile matrix generated from scRNA-seq data to deconvolute GeoMx® data. Danaher *et al*. (2020) which contains 18 cell types was used for this analysis ^41^.

Pathway analysis was performed to explore the different aggregate gene sets. Each individual ROI/AOI segment was scored for every pathway of interest, which we then used to investigate biological differences. Pathways and gene sets were defined from the Reactome database (https://reactome.org/). We used the R software package called Gene Set Variation Analysis to score each segment within our study (https://www.bioconductor.org/packages/release/bioc/html/GSVA.html). A total of 1,557 gene sets were scored (we calculated the scores for any gene set with at least 5 genes and fewer than 500 genes).

### 2.6. Data Analysis and Differential Expression Analysis Using GeoMx® DSP

Statistical analysis was performed using R software (R Core Team, 2023). Differential expression (DE) analysis and gene set enrichment analysis (GSEA) were performed using data generated from the NanoString GeoMx® DSP platform. ROIs were segmented by phenotype (PanCK+, S100+, and CD45+) and analyzed for gene expression differences across the three diagnostic groups (TC, PG, and PPCT). Raw counts were normalized using Q3 normalization followed by log2 transformation, as implemented in the GeoMx® NGS pipeline ^42^.

Differential gene expression was assessed using the linear modeling framework of the limma-voom method ^43^, which applies precision weights to log2-transformed expression values and fits a linear model with the diagnostic group as the main covariate. For each gene, an empirical Bayes moderated *t*-statistic was computed to derive the unadjusted *p*-value. To account for multiple testing, the resulting *p*-values were adjusted using the Benjamini–Hochberg (BH) method ^44^ to control the false discovery rate (FDR). Genes were considered statistically significant to have robust DE at FDR < 0.05.

Statistical analyses and report generation were performed using the GeoMx® DSP Analysis Suite (NanoString Technologies, Inc.) and supplemented with downstream visualization in R (v4.3.1) using limma, edgeR, and ggplot2 packages. Some figures and graphs were generated using GraphPad Prism (version 10.5.0, GraphPad Software, San Diego, CA).

## 3. Results

### 3.1. Patient Demographics and Tumor Types

The cohort included 38 patients with a median age of 61.5 years (range 19 – 78 years; IQR 49 – 68 years), 76% of whom were females. TC and PPCT presented in the lung, while PG were extra-adrenal. Most patients were non-smokers (73.6%), and the majority were Hispanic whites (55.2%). Tumor sizes ranged from 0.5 cm to 4.9 cm (median 2.0 cm; IQR 1.025 – 2.500), with TC generally smaller than PG. Pathologic staging data indicated that most TC and PPCT were early-stage (pT1a – pT2b, pN0) (**Supplementary Table S1**).

The 38 tumors from 38 patients consisted of 20 TC (65% females; median age: 63.5 years), 16 PG (87.5% females; median age: 52.5 years), and 2 PPCT (100% females; ages 66 and 70 years). All tumors showed neuroendocrine morphology on H&E. PPCT displayed granular dark-brown cytoplasmic pigment and a nested (zellballen) architecture.

Of the 38 cases, all were profiled by IHC (**Figure 2**), and 36 were included in the spatial transcriptomic analysis via NanoString GeoMx® DSP. The analysis produced 74 high-quality ROIs that enabled transcriptomic and immune profiling across all tumor subtypes.

**Figure 2.**
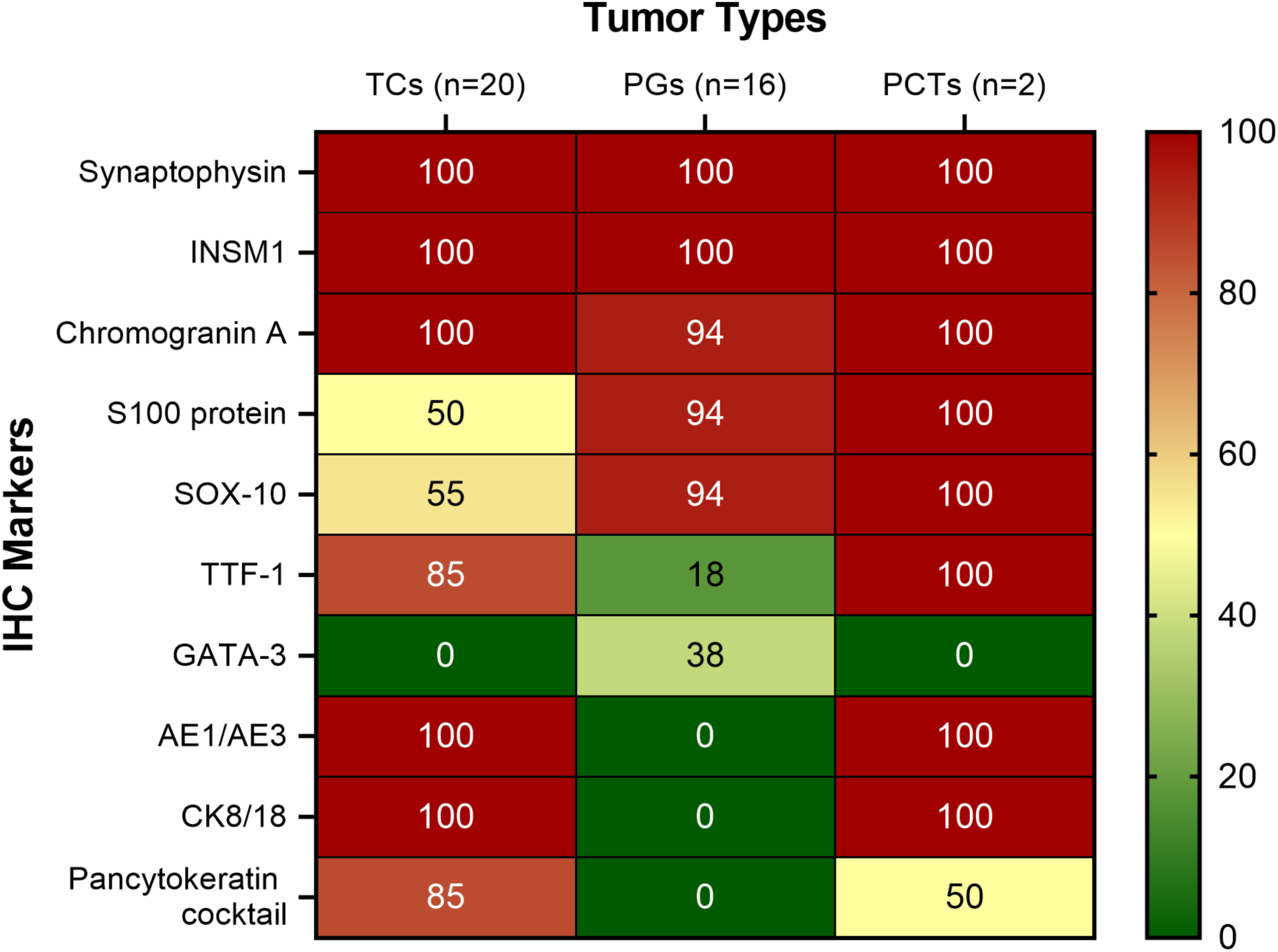
Heatmap of immunohistochemical marker expression across the tumor types. This heatmap illustrates the percentages of positive IHC staining for 10 diagnostic markers across the three tumor categories: TC (n = 20), PG (n = 16), and PPCT (n = 2). Markers include neuroendocrine markers (Synaptophysin, INSM1, Chromogranin A), neural crest/sustentacular markers (S100 protein and SOX-10), pulmonary epithelial markers (TTF-1), GATA-3, and epithelial cytokeratins (AE1/AE3, CK8/18, and PanCK cocktail). Color intensity reflects the proportion of tumors staining positive for each marker, with red indicating high expression (100%) and green indicating low or absent expression (0%). PPCT display a hybrid immunophenotype, overlapping with both TC (shared neuroendocrine and epithelial markers) and PG (S100 and SOX-10 positivity).

### 3.2. Immunohistochemical Profiling

Synaptophysin and INSM1 were diffusely positive in 100% of cases across all tumor types. Chromogranin A was diffusely positive in 100% of TC and PPCT, and in 94% (15/16) of PG. S100 positivity was observed in 50% of TC (10/20), 94% of PG (15/16), and 100% of PPCT. SOX-10 showed a similar distribution, with positivity in 55% of TC (11/20), 94% of PG (15/16), and 100% of PPCT. TTF-1 was expressed in 85% of TC (17/20), 100% of PPCT, and 18% of PG (3/16). GATA-3 was negative in all TC and PPCT but positive in 38% of PG (6/16) (**Supplementary Table S2; Figure 2**). Cytokeratin expression patterns were as follows: cytokeratins AE1/AE3 and CK8/18 were diffusely positive in all TC and both PPCT cases but negative in all PG. PanCK cocktail was positive in 85% of TC (17/20), 50% of PPCT (1/2), and 0% of PG (**Supplementary Table S2; Figure 2**).

Most tumors exhibited a low Ki-67 index (<5%). One case of TC demonstrated an intermediate Ki-67 labeling index of 11 – 20%, while one PG exhibited a higher proliferation index (31 – 40%). Both PPCT had low Ki-67 indices (<5%) (**Supplementary Table S2**).

### 3.3. GeoMx® DSP Spatial Transcriptomics Profiling

Spatially resolved ROIs were defined based on PanCK, S100, and CD45 immunofluorescence stains and classified into three categories: PanCK+ ROIs: epithelial-rich compartments; S100+ ROIs: neural crest/sustentacular-rich regions; CD45+ ROIs: immune cell-rich areas (**Figure 1**; **Supplementary Fig. S2**). A total of 304 ROIs were profiled, distributed across representative cases of each tumor subtype.

Uniform Manifold Approximation and Projection (UMAP) analysis of 74 spatial transcriptomic ROIs revealed distinct clustering by diagnostic group. PG formed a discrete cluster, while TC also clustered separately. Notably, PPCT were embedded within the TC cluster, suggesting greater transcriptional similarity to carcinoids, as expected. Segment-level annotations revealed epithelial and sustentacular contributions to the clustering patterns, with PanCK+ and S100+ segments differentially enriched in TC, PG and PPCT (**Figure 3**).

**Figure 3.**
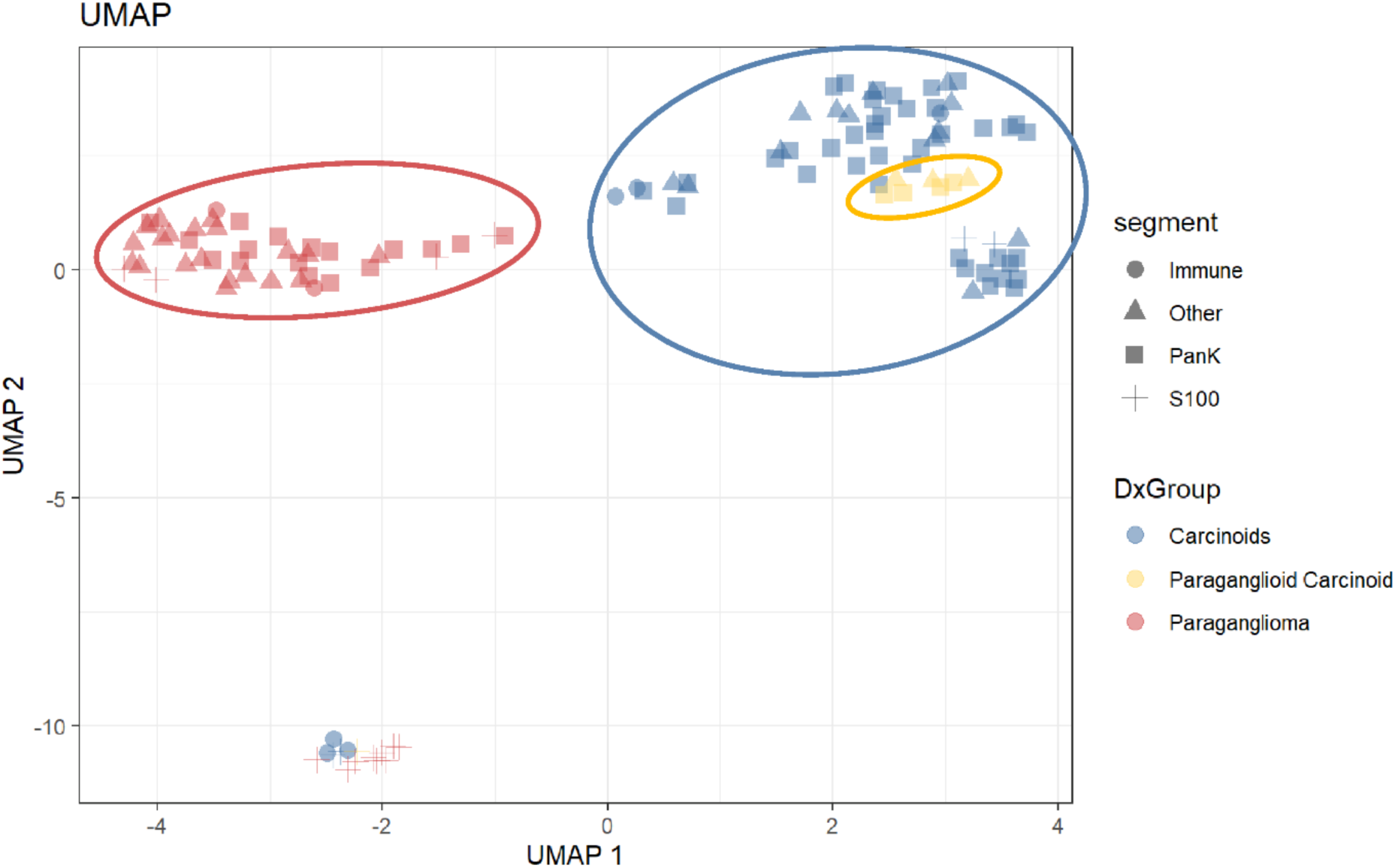
UMAP clustering of spatial transcriptomic ROIs by diagnostic group and phenotypic segment. Uniform Manifold Approximation and Projection (UMAP) analysis of 74 NanoString GeoMx® DSP ROIs demonstrates distinct clustering by diagnosis. PG (red) form a discrete cluster, while TC (blue) are separately grouped. PPCT (yellow) cluster within the TC group, indicating closer transcriptional similarity to carcinoids. Each point represents an ROI and is shaped by segment phenotype: PanCK+ (epithelial), S100+ (sustentacular), CD45+ (immune), or other. The spatial organization suggests that both diagnostic category and segment phenotype contribute to the observed transcriptomic variance.

### 3.4. Differential Gene Expression Transcriptomic Profiling

We performed a comparative transcriptomic analysis across four distinct contrasts involving PG, PPCT, and TC using GeoMx® DSP data (**Figure 1**; **Table 1**). Each contrast provided insight into the molecular landscape distinguishing these neuroendocrine tumors. We employed differential expression analysis on a per-gene basis where the normalized expression is modeled using a linear mixed-effect model (LMM) to account for the sampling of multiple ROI/AOI segments per tissue. Differential expression analysis was performed across four contrasts:

**Table 1.**
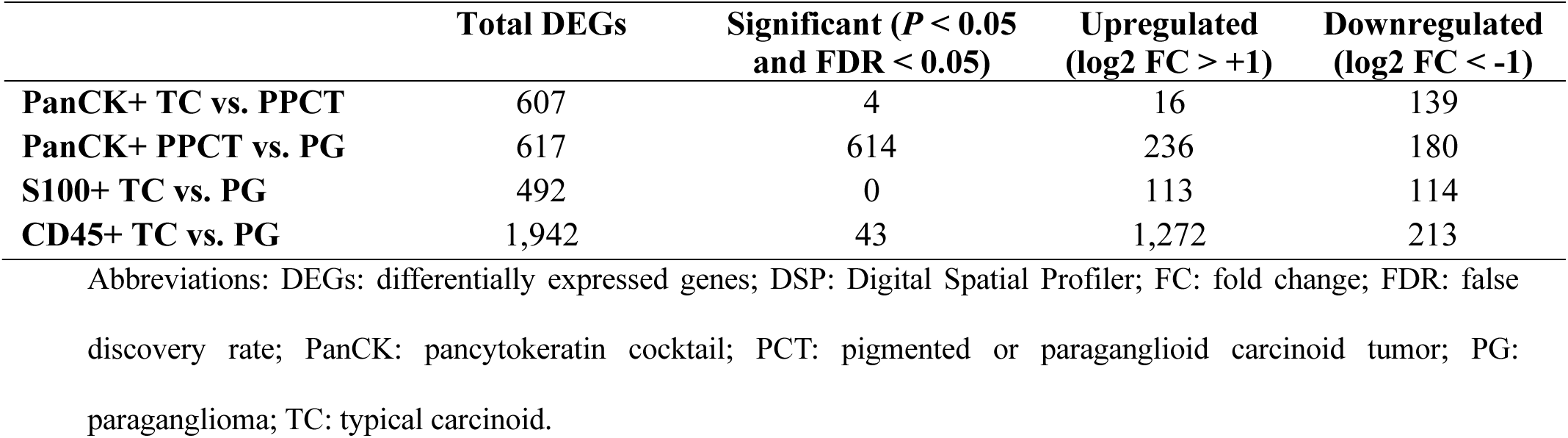
Summary of the differential gene expression analysis across the three diagnostic groups and segmented compartments. This table summarizes the number of DEGs identified across selected diagnostic group comparisons in spatial transcriptomic analysis using the GeoMx® DSP. Analyses were stratified by segmented compartments based on marker expression (PanCK+, S100+, CD45+). Total DEGs were determined based on nominal *p*-value < 0.05, with significance defined as both *p* < 0.05 and FDR < 0.05. Genes were further classified as upregulated (log2 FC > +1) or downregulated (log2 FC < −1) in the respective comparison.

#### 3.4.1. PanCK+ PPCT vs. TC

The PanCK compartment yielded 607 evaluable genes, of which 139 genes were downregulated (using the thresholding of log2 FC < –1) and 16 were upregulated (using the thresholding of log2 fold change > +1), but only 4 genes reached statistical significance (*P* < 0.05 and FDR < 0.05) and are *CST2*, *CST4*, *FBXO22*, and *DPP8* (**Figure 4**). The most significant gene was *CST2*, showing strong upregulation (log2 FC = 5.58, FDR = 0.0049) in PPCT relative to TC. *CST2* encodes cystatin SA, a cysteine protease inhibitor potentially implicated in tumor immune evasion. A heatmap of these 607 genes shows clustering by the diagnosis group (**Supplementary Table S3; Supplementary Fig. S3**).

**Figure 4.**
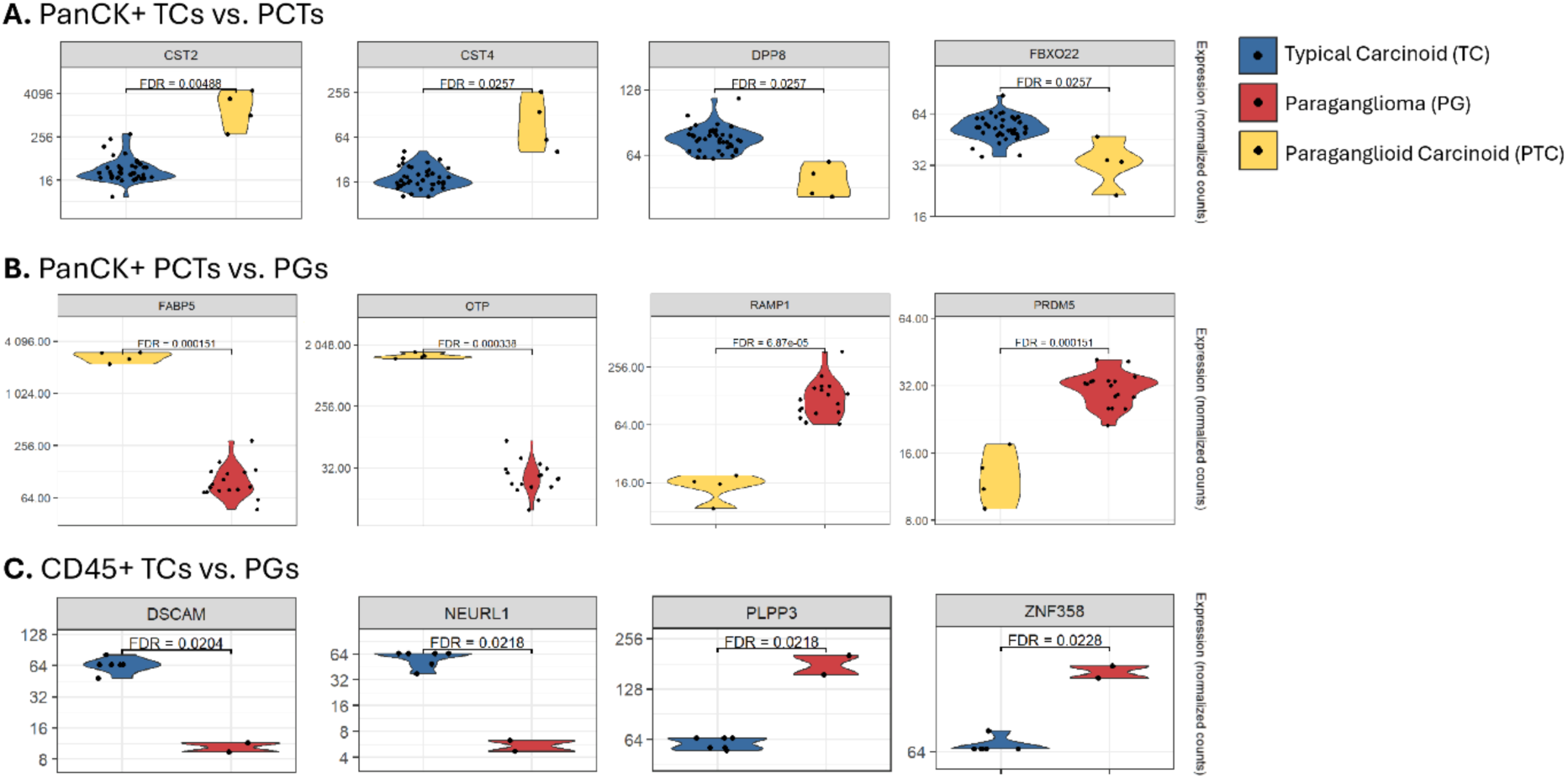
Differential gene expression across TC, PPCT, and PG based on cell phenotype and diagnostic group. Violin plots depict normalized gene expression counts for selected significant differentially expressed genes across three comparison groups, with false discovery rate (FDR) indicated above each plot. **(A)** PanCK+ TC vs. PPCT: comparison of epithelial (PanCK positive) typical carcinoids (blue) and paraganglioid carcinoid tumors (yellow). Genes such as *CST2* and *CST4* are significantly upregulated in PPCT relative to TC, whereas *DPP8* and *FBXO22* are significantly downregulated in PPCT relative to TC. **(B)** PanCK+ PPCT vs. PG: comparison of epithelial (PanCK positive) paraganglioid carcinoid tumors (yellow) and paragangliomas (red). Genes such as *FABP5* and *OTP* are significantly upregulated in PPCT relative to PG, whereas *RAMP1* and *PRDM5* are enriched in PG relative to PPCT. **(C)** CD45⁺ TC vs. PG: comparison of immune-rich CD45+ fractions of typical carcinoids (blue) versus paragangliomas (red). *DSCAM* and *NEURL1* are significantly overexpressed in TC relative to PG, while *PLPP3* and *ZNF358* are significantly upregulated in PG relative to TC.

#### 3.4.2. PanCK+ PPCT vs. PG

This comparison yielded the highest number of differentially expressed genes with 614 out of 617 genes showing *P* < 0.05 and FDR < 0.05. *OTP*, *DIRAS3*, *CST2*, *MS4A8*, and *FABP5* were among the top upregulated genes in PPCT relative to PG, while *OLFM1*, *RAMP1*, *NDUFA4L2*, *PHOX2A*, and *PRDM5* were among the top downregulated genes in PPCT relative to PG (**Figure 4**). A heatmap of these genes shows clustering by the diagnosis group (**Supplementary Table S4; Supplementary Fig. S4**).

#### 3.4.3. S100+ TC vs. PG

Out of 492 evaluated genes in the S100+ compartment, 113 genes were upregulated using the thresholding of log2 FC > +1 and 114 genes were downregulated using the thresholding of log2 FC < –1; however, none reached FDR significance (*P* < 0.05 and FDR < 0.05). A heatmap of the 492 genes shows clustering by the diagnosis group (**Supplementary Table S5; Supplementary Fig. S5**).

#### 3.4.4. CD45+ TC vs. PG

In the immune compartment, 1,272 out of the 1,942 genes were upregulated using the thresholding of log2 FC > +1 and 213 were downregulated using the thresholding of log2 FC < –1. A total of 43 genes reached FDR significance (*P* < 0.05 and FDR < 0.05). Interestingly, *NEURL1*, *DSCAM*, *RNF122*, *H1-5*, and *KAT6A* were among the top upregulated genes in TC relative to PG, whereas *PANK3*, *RABAC1*, *PLPP3*, *TBC1D4*, and *ZNF358* were among the top downregulated genes in TC relative to PG (**Figure 4**). A heatmap of the 1,942 genes shows clustering by the diagnosis group (**Supplementary Table S6; Supplementary Fig. S6**).

#### 3.4.5. Differential Expression of Melanocytic Differentiation and Neural Crest Markers in PPCT vs. TC

To investigate whether PPCT preferentially express markers indicative of partial melanocytic or neural crest differentiation, which may underlie their morphological distinctions from TC, we conducted a targeted differential gene expression analysis focusing on established melanocyte-associated and neural crest lineage markers. Among the genes significantly upregulated in PPCT relative to TC, several markers of melanocytic differentiation were noted, including *S100B* (Log2 FC = 1.725, FDR = 0.69) and *GPNMB* (Log2 FC = 1.634, FDR = 0.53), both previously implicated in pigment cell differentiation and melanoma biology ^45–48^. Additionally, *FABP5*, which encodes for a fatty acid-binding protein associated with melanogenesis and metastatic melanoma progression, demonstrated marked upregulation in PPCT relative to TC (Log2 FC = 3.16, FDR = 0.64) (**Figure 5A**).

**Figure 5.**
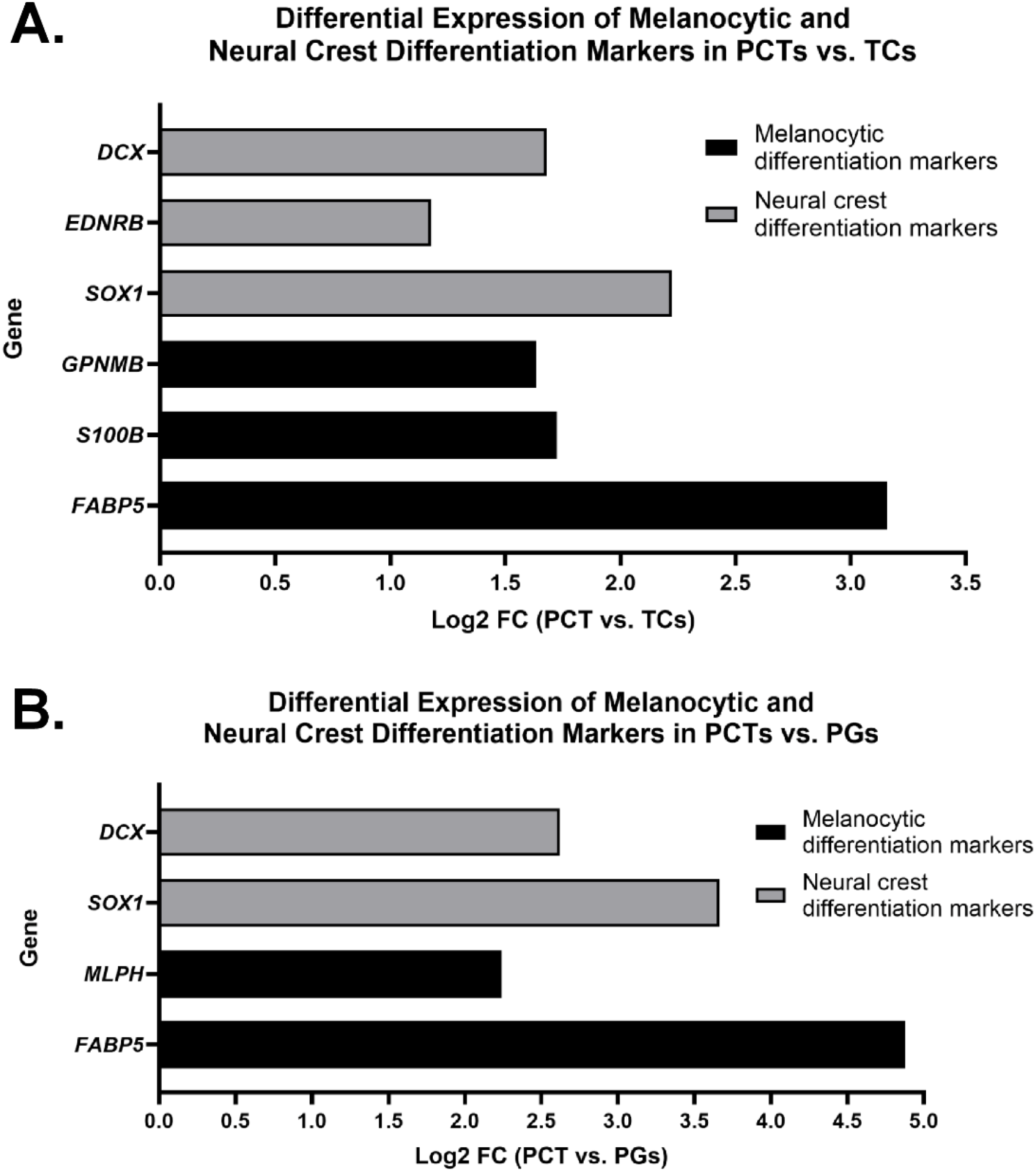
Differential expression of melanocytic and neural crest differentiation markers in paraganglioid carcinoid tumors (PPCT) vs. typical carcinoids (TC) and paragangliomas (PG). **(A)** Bar graph showing log2 fold change (Log2 FC) of selected genes upregulated in PPCT relative to TC, based on transcriptomic analysis. Genes associated with melanocytic differentiation (black bars) include *FABP5*, *S100B*, and *GPNMB*, while genes of neural crest lineage (gray bars) include *SOX1*, *EDNRB*, and *DCX*. **(B)** Bar graph showing Log2 FC of the same gene categories in PPCT compared to PG. Melanocytic differentiation markers (black bars) include *FABP5* and *MLPH*, whereas neural crest differentiation markers (gray bars) include *SOX1* and *DCX*. The upregulation of these genes in PPCT supports partial activation of melanocytic and neural crest differentiation programs in this histologic variant. Positive Log2 FC values indicate relative overexpression in PPCT.

Genes associated with neural crest development and migration, including *SOX1* (Log2 FC = 2.221, FDR = 0.77) ^37,49^, *EDNRB* (Log2 FC = 1.177, FDR = 0.53) ^50,51^, and *DCX* (Log2 FC = 1.68, FDR = 0.76) ^52^, also exhibited upregulation in PPCT vs. TC, suggesting higher activation of neural crest-related transcriptional programs in PPCT (**Figure 5A**). However, canonical melanocyte-lineage defining markers such as *TYR*, *DCT*, and *MITF* were not detected ^53,54^, indicating that the observed differentiation may be partial or incomplete. Also, despite the lack of statistical significance by FDR, biological plausibility remains relevant, taking into account the large log2 FCs warranting further validation.

#### 3.4.6. Differential Expression of Melanocytic Differentiation and Neural Crest Markers in PPCT vs. PG

Transcriptomic analysis comparing PPCT to PG revealed significant differential expression of genes associated with melanocytic and neural crest lineages. Among melanocyte-associated genes, *FABP5* (Log2 FC = 4.878, FDR = 0.00015) and *MLPH* (Log2 FC = 2.240, FDR = 0.0094) were markedly upregulated in PPCT. Similarly, neural crest differentiation markers such as *SOX1* (Log2 FC = 3.662, FDR = 0.00099) and *DCX* (Log2 FC = 2.621, FDR = 0.016) also demonstrated significant upregulation in PPCT compared to PG (**Figure 5B**). These findings support the hypothesis that PPCT exhibit partial melanocytic differentiation while maintaining a neural crest-associated transcriptional program.

### 3.5. Gene Set and Pathway Enrichment Analysis

Using the statistical criteria FDR < 0.2 and *p* < 0.05, comparative gene set enrichment and pathways analyses highlighted key transcriptomic distinctions across the epithelial (PanCK+), neural crest/sustentacular-rich (S100+), and immune-rich (CD45+) compartments among PPCT, TC, and PG. Gene set enrichment analysis (GSEA) revealed nine differentially enriched pathways in PPCT relative to TC. Among these, several pro-inflammatory and stress-response pathways were significantly upregulated in PPCT, including the *TRAF6*-mediated induction of the *TAK1* complex within the *TLR4* signaling cascade (Log2 FC = 1.06, FDR = 0.023). Notably, the regulation of *PTEN* localization (Log2 FC = 1.43, FDR = 0.096) and inositol phosphate metabolism (Log2 FC = 1.73, FDR = 0.11) pathways were enriched (**Figure 6 A**), implicating roles in membrane signaling and intracellular trafficking which may be potentially relevant to melanosome biology. Only one pathway, polo-like kinase (PLK)– mediated events, was significantly downregulated in PPCT compared to TC (Log2 FC = –1.67, FDR = 0.15; **Figure 6B**) (**Supplementary Table S7**; **Supplementary Fig. S7**).

**Figure 6.**
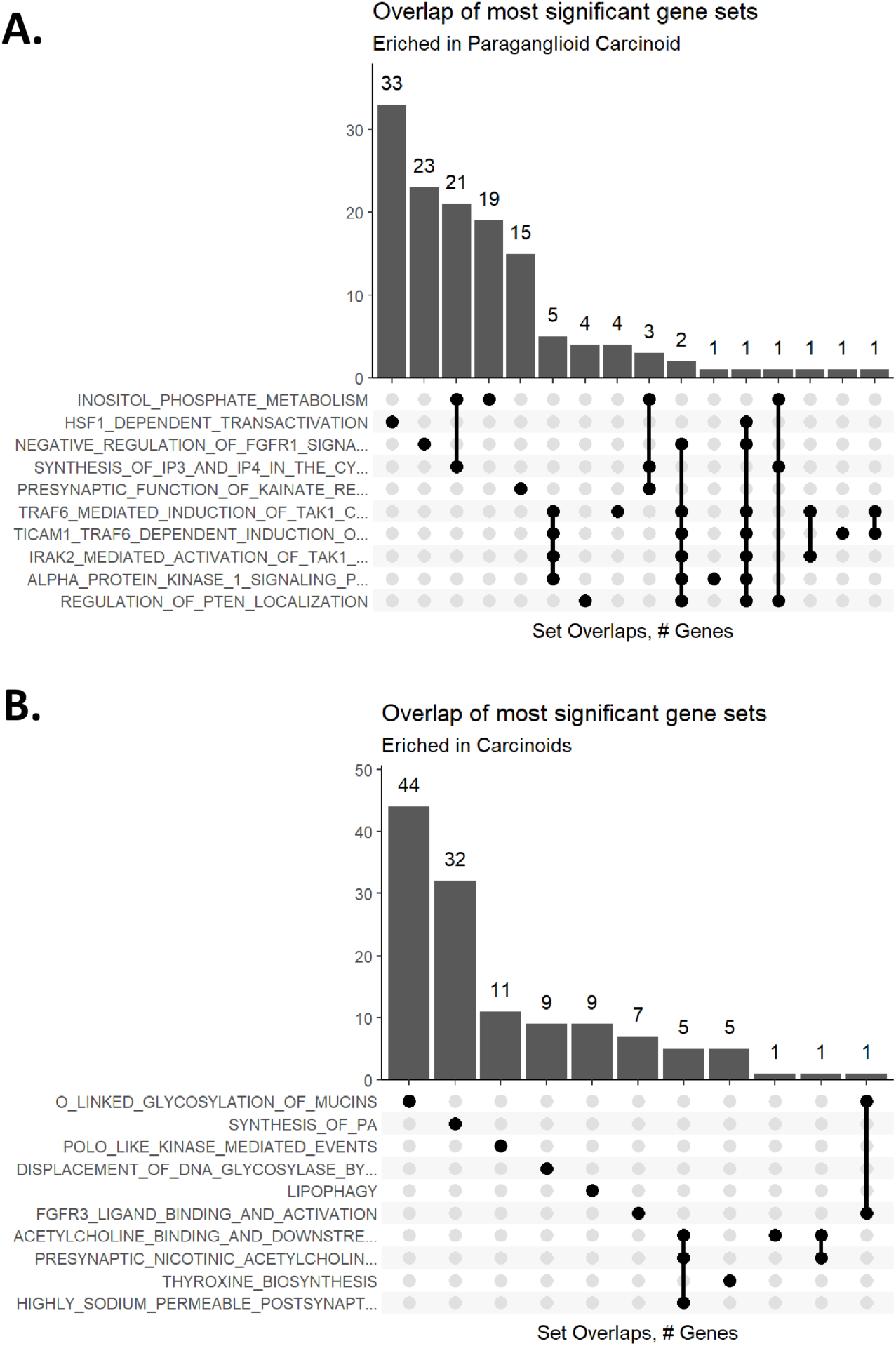
Selected pathway overlap and transcriptional reprogramming in PanCK+ compartments. **(A)** UpSet plot highlighting the most enriched gene sets in PPCT. **(B)** UpSet plot highlighting gene sets highly enriched in TC.

A total of 289 enriched pathways were identified in PanCK+ PPCT vs. PG (**Supplementary Table S8**; **Supplementary Fig. S8**), underscoring substantial molecular divergence driven by the epithelial tumor cells. Among these, upregulated pathways included *NEF*-mediated downregulation of MHC class I and voltage-gated potassium channels, whereas keratan sulfate biosynthesis and ephrin-mediated cell repulsion were markedly downregulated. The S100+ Schwann-like tumor cell-enriched TC vs. PG exhibited minimal divergence, with only 4 enriched pathways, including modest upregulation of HIV-1 *NEF*–related signaling and downregulation of kainate receptor–mediated neurotransmission (**Supplementary Table S9**; **Supplementary Fig. S9**). The CD45+ immune cell-rich compartment showed 93 enriched pathways (**Supplementary Table S10**; **Supplementary Fig. S10**), including high upregulation of γ-carboxylation/hypusinylation pathways and amine-derived hormone metabolism, while pathways such as ADP signaling via P2Y receptors and *L1CAM* interactions were strongly suppressed. These findings show that the epithelial compartment (PanCK+) may be the primary driver of the unique transcriptional identity of PPCT compared to PG and TC.

## Discussion

PPCT of the lung represent a rare and often underrecognized subgroup of pulmonary NETs with notable clinical and pathologic similarities to extra-adrenal PG ^11–16,18,19^. We performed comprehensive analysis of PPCT by comparing them to TC and PG using histopathologic, immunohistochemical, and digital spatial transcriptomic profiling. Our results indicate that the transcriptomic characteristics of PPCT are consistent with those of TC and distinct from PG, as anticipated; however, the upregulated expression of melanocytic and neural crest differentiation markers may offer a molecular basis for the morphological differences observed between PPCT and TC and the similarities to PG.

The hallmark of neuroendocrine differentiation in pulmonary tumors is the expression of markers such as synaptophysin, chromogranin A, and INSM1 ^34,55,56^. Our data confirmed the uniform expression of these neuroendocrine markers across all three tumor types, including PPCT, consistent with prior reports ^11–13^. However, PPCT also demonstrated robust S100 and SOX-10 expression in a pattern similar to that of the sustentacular cells of PG. TC and PPCT consistently demonstrated cytokeratin AE1/AE3 and CK8/18 expression by IHC, whereas PG did not exhibit any epithelial markers. Notably, one PPCT in our cohort showed no significant expression of PanCK, indicating that a loss of epithelial marker expression may occur in PPCT. This aligns with rare reports suggesting that lung carcinoids can occasionally be cytokeratin-negative, particularly in tumors located in the periphery^56^. While the distinction between these rare TC from PG may be clinically insignificant, these may represent a diagnostic pitfall^29,30,32,33^. We recommend the use of AE1/AE3, CK8/18 or more than one cytokeratin to avoid this pitfall.

TTF-1, a nuclear transcription factor typically expressed in pulmonary epithelial and neuroendocrine cells, was retained in all PPCT and most TC but was largely absent in PG. This marker, therefore, remains a critical discriminator between pulmonary-origin NETs and extra-pulmonary NETs ^34,57–59^. Conversely, GATA-3, a transcription factor more commonly expressed in PG and various endocrine tissues ^60–62^, was expressed in 38% of PG but not in PPCT or TC.

Beyond conventional histopathology, we used the NanoString GeoMx® DSP platform to gain deeper insight into the transcriptomic landscape of PPCT. DSP enables spatially resolved transcriptomic profiling by allowing for the quantification of RNA within precisely defined tissue compartments ^63,64^. By selecting ROIs based on immunofluorescent labeling of PanCK, CD45 and S100, we interrogated epithelial (PanCK+), neural crest-derived/sustentacular-rich (S100+), and immune cell-rich (CD45+) regions within our tumor samples.

Our transcriptomic analysis showed that PPCT exhibit enriched expression of melanocytic as well as neural crest markers, including upregulation of *FABP5* – a fatty acid-binding protein involved in melanin metabolism and melanoma biology ^65^, *MLPH* (melanophilin) – which plays a role in melanosome transport ^66^, and *S100B* and *GPNMB* – both which had been previously implicated in pigment cell differentiation and melanoma biology ^45–48^. Additionally, *SOX1*, *EDNRB*, and *DCX*, which are markers of neural progenitors and neural crest-derived lineages ^37,49–52^ were significantly enriched in PPCT. Our transcriptomic findings may also explain the observed S100+ sustentacular cell network in PPCT and neuromelanin pigmentation seen in these tumors.

The gene set and pathway enrichment analyses demonstrated selective upregulation of pro-inflammatory and stress-response signaling pathways, such as the *TRAF6*-mediated activation of the *TAK1* complex within the *TLR4* cascade, implicating innate immune activation and inflammatory crosstalk in the epithelial tumor niche ^67,68^. Additionally, the enrichment of pathways regulating *PTEN* localization and inositol phosphate metabolism suggests altered membrane trafficking and phosphoinositide signaling ^69^, which may be mechanistically linked to intracellular organelle dynamics, including melanosome transport and function ^70^, an observation potentially aligned with the partial melanocytic differentiation phenotype of PPCT. Conversely, the downregulation of polo-like kinase (*PLK*)-associated pathways may reflect altered cell cycle control and mitotic regulation in these tumors ^71,72^.

A primary limitation of our study is the small number of PPCT cases. Although we attempted to address this by employing duplicate punches in the TMA with multiple ROIs, further research is necessary to validate our results.

## Conclusion

This study offers a detailed molecular and spatial profile of PPCT of the lung, integrating IHC and spatial transcriptomic profiling. PPCT display gene signatures similar to TC with unique upregulation of melanocytic and neural crest lineages and pathways related to inflammation, membrane trafficking, and melanosome-related signaling. These results may partly explain the morphologic resemblance to PG.

## Author Contributions

H.F.B., L.M.B., and R.R.-C. were involved in the study design and writing. A.PT.-T, B.M.C.-G., R.S.-V., J.A.-L., K.D.-E., L.M.B., M.L.-G., and R.R.-C. were involved in sample contribution and data collection. H.F.B., A.R, A.P., L.M.B. and R.R.-C. contributed to data analysis. H.F.B., L.M.B. and R.R.-C. were involved in the development of methodology. All authors read and approved the final manuscript.

## Data Availability Statement

The original contributions presented in this work are included in the article. Further inquiries can be directed to the senior author.

## Ethics Approval and Consent to Participate

The study was approved by the Institutional Review Board of the University of Miami Miller School of Medicine. All data were handled in compliance with the ethical standards of the Declaration of Helsinki. Patient identifiers were removed to ensure confidentiality, and only de-identified data were used in analysis. The study posed no direct risk to patients.

## Acknowledgments

Statistical Analyses were performed in collaboration with Bruker Spatial Biology. Figure 1 was created with BioRender.com (accessed in July 2025). All rights and ownership of BioRender content are reserved by BioRender. BioRender content included in the completed graphic is not licensed for any commercial uses beyond publication in a journal. For any commercial use of this figure, users may, if allowed, recreate it in BioRender under an Industry BioRender Plan.

## Supplementary Figures

**Supplementary Figure S1.**
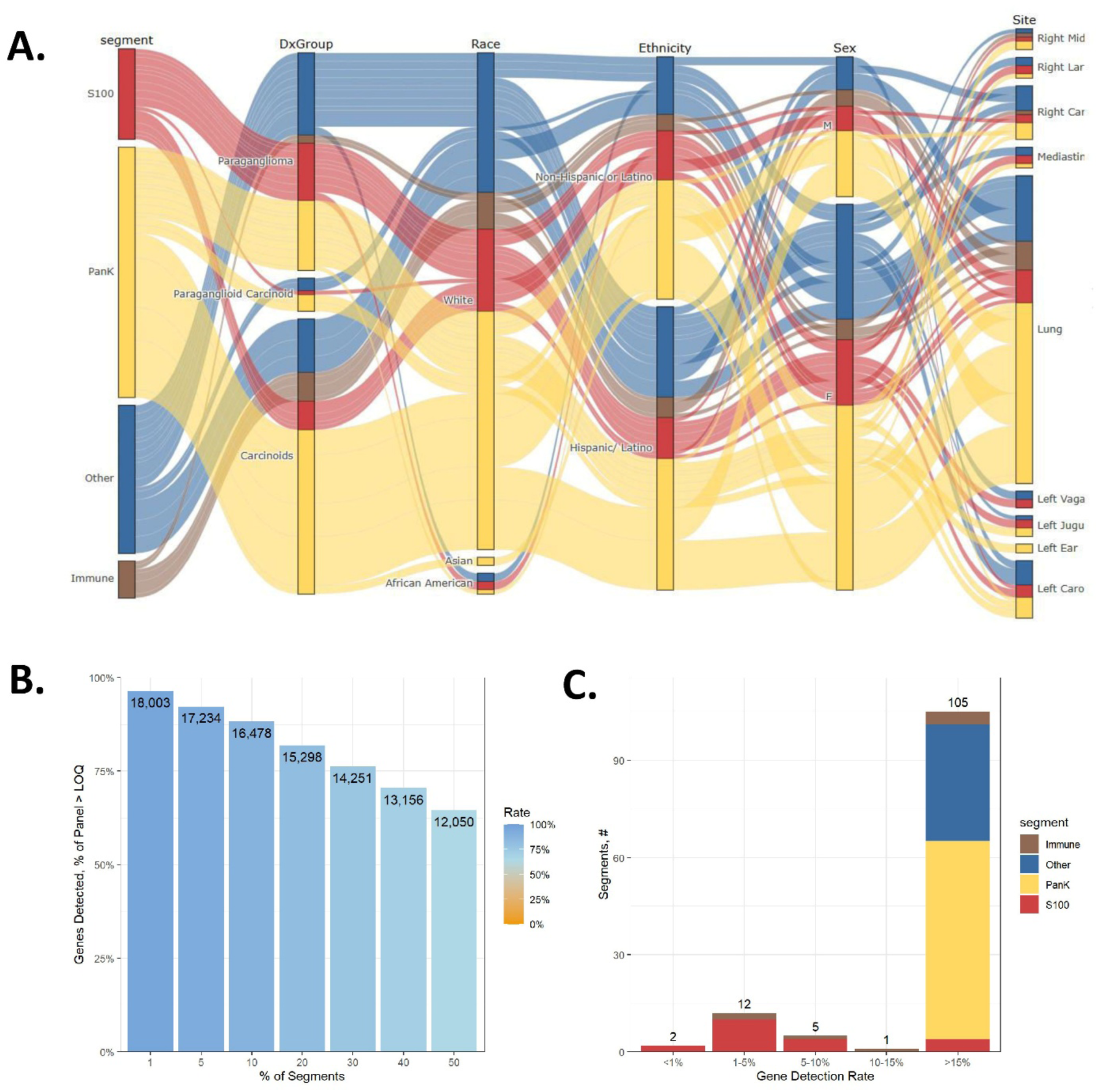
Overview of the study design and quality control (QC) metrics. **(A)** Sankey diagram summarizing the study design. Each column represents biological or demographic annotations (segment type, diagnosis group, race, ethnicity, sex, and anatomic site), with flows (cords) visualizing how individual ROI/AOI segments relate across variables. Five segments were excluded due to QC failures or low gene detection rates. **(B)** Bar plot showing the number of genes detected above the lower limit of quantification (LOQ) across increasing proportions of segments. A total of 14,251 genes detected in at least 30% of segments, were explored in depth. **(C)** Distribution of gene detection rates across segments by annotation group. The majority of PanCK+ and S100+ segments showed >16% detection rates, while a few immune segments had detection rates below 10%.

**Supplementary Figure S2.**
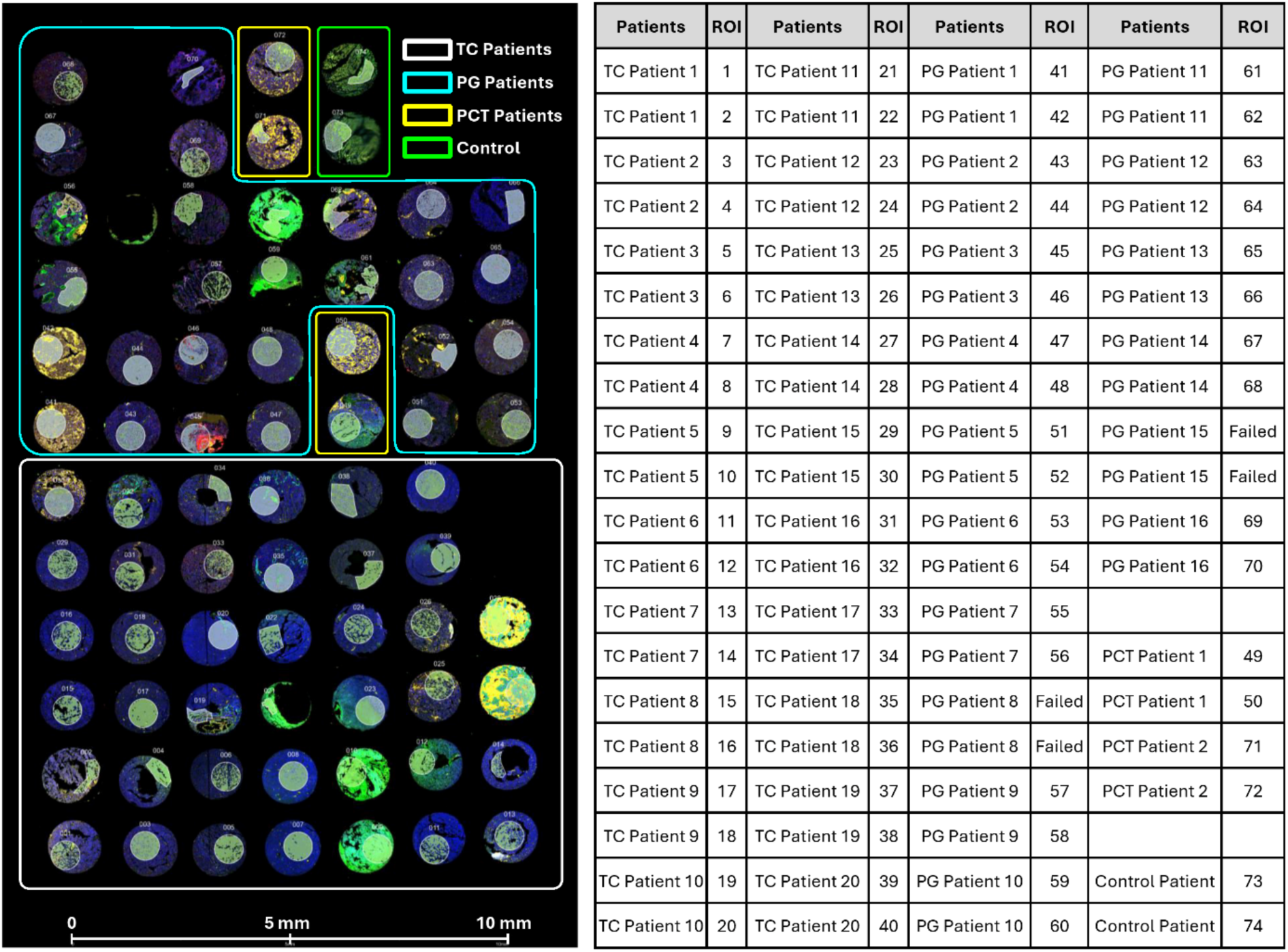
Digital spatial profiling (DSP) ROI map of the tested tumors. Representative fluorescent image of the tissue microarray (TMA) used for NanoString GeoMx® DSP analysis, showing all 76 tissue cores included in the analysis. Each circular core corresponds to a case of TC (n = 20), PG (n = 14), or PCT (n = 2), color-coded by patient group (white = TC, cyan = PG, yellow = PPCT, green = control patient). Regions of interest (ROIs) were selected within each core based on immunofluorescent staining for epithelial and neural crest markers: blue = DAPI (nuclear stain), green = PanCK, red = S100, yellow = CD45. The bar at the bottom denotes a spatial scale of 0 – 10 mm. The table on the right summarizes the patient identifiers, diagnostic group, and corresponding ROI numbers (1 – 74). ROIs marked as “Failed” indicate unsuccessful data acquisition due to quality control issues (PG patients 8 and 15).

**Supplementary Figure S3.**
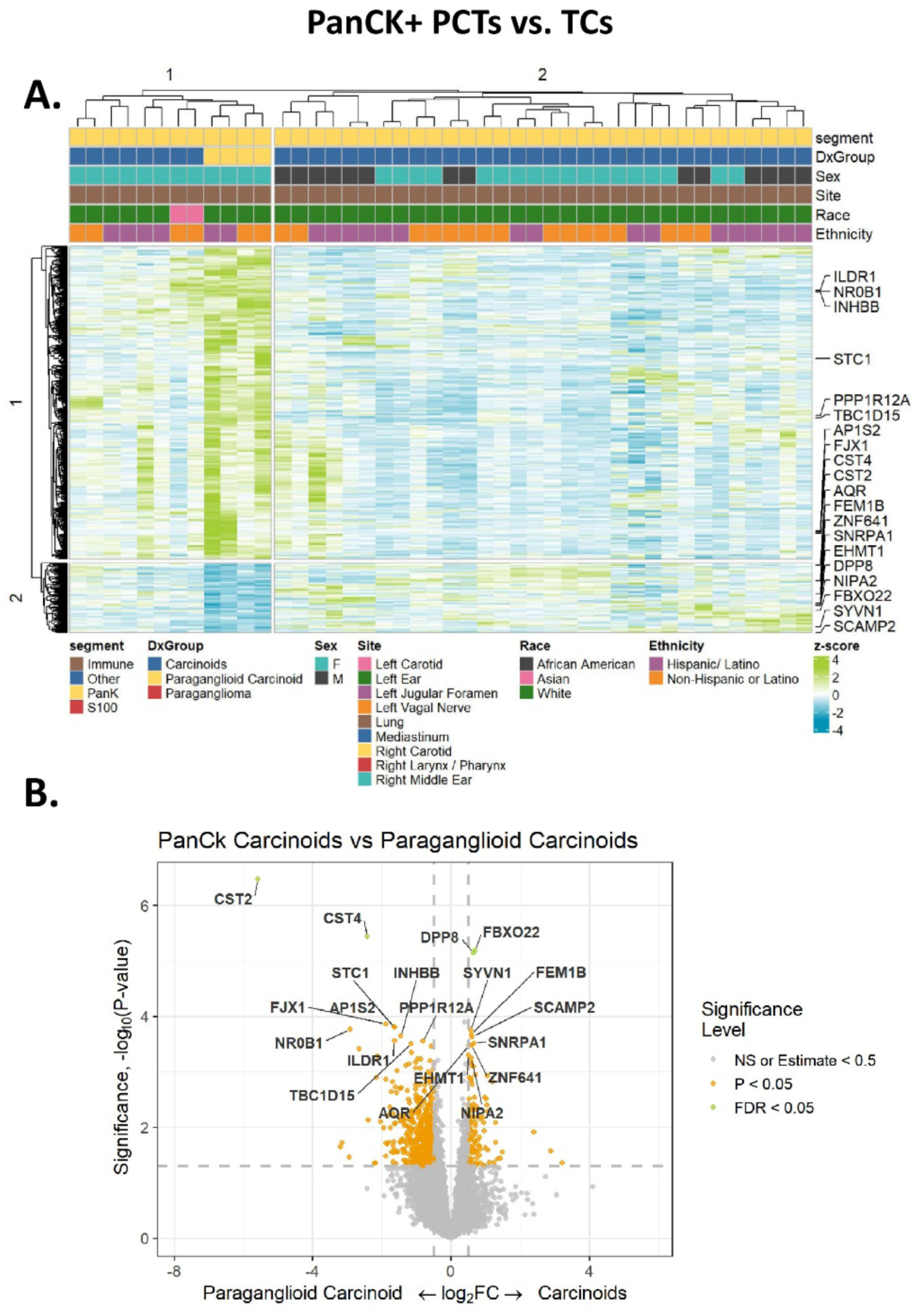
Heat map and volcano plot of the differential gene expression profiles between PanCK+ PPCT vs. TC. **(A)** Heatmap displaying the expression of differentially expressed genes between PanCK-positive PPCT and TC. Columns correspond to PanCK+ ROIs, annotated by diagnosis, tissue site, and demographics. **(B)** Volcano plot depicting differential gene expression between PanCK-positive TC and PPCT. The x-axis represents the log2-transformed fold change (log2 FC), with positive values indicating higher expression in TC and negative values indicating higher expression in PPCT. The y-axis represents the –log10-transformed *p*-values, reflecting statistical significance. Genes with a false discovery rate (FDR) threshold of 0.05 are highlighted in green, nominally significant genes (*p* < 0.05) are in orange, and non-significant genes or those with minimal fold change (|log2 FC| < 0.5) are shown in grey. Genes significantly upregulated in PPCT relative to TC include *CST2* and *CST4*.

**Supplementary Figure S4.**
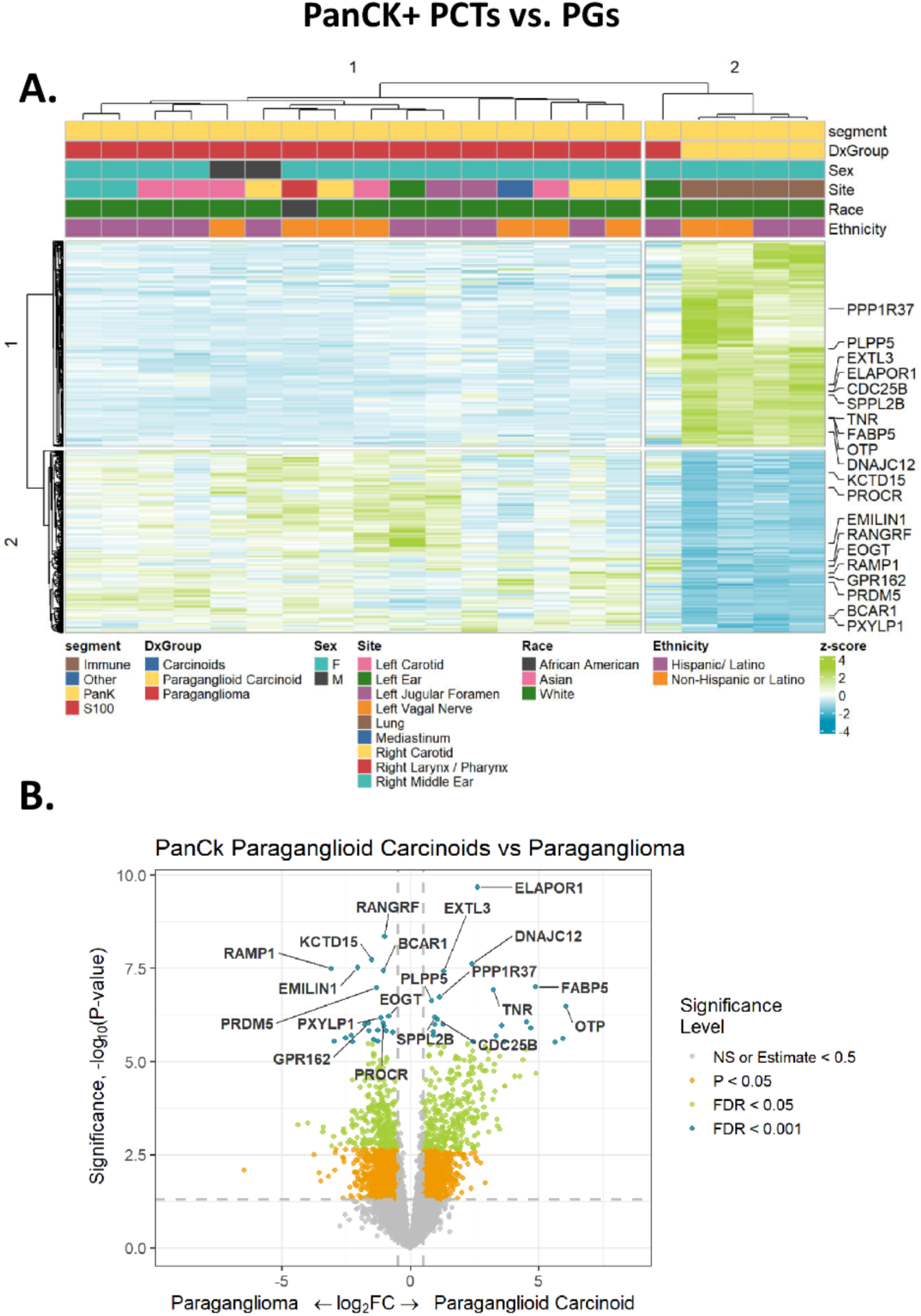
Heat map and volcano plot of the differential gene expression profiles between PanCK+ PPCT vs. PG. **(A)** Heatmap displaying the expression of differentially expressed genes between PanCK-positive PPCT and PG. Columns correspond to PanCK+ ROIs, annotated by diagnosis, tissue site, and demographics. **(B)** Volcano plot depicting differential gene expression between PanCK-positive PPCT and PG. The x-axis represents the log2-transformed fold change (log2 FC), with positive values indicating higher expression in PPCT and negative values indicating higher expression in PG. The y-axis represents the –log10-transformed *p*-values, reflecting statistical significance. Genes with a false discovery rate (FDR) threshold of 0.05 are highlighted in green, those with FDR threshold of 0.001 are highlighted in blue, nominally significant genes (*p* < 0.05) are in orange, and non-significant genes or those with minimal fold change (|log2 FC| < 0.5) are shown in grey. Genes such as *ELAPOR1*, *FABP5*, and *OTP* demonstrate strong differential expression favoring PPCT, while *RAMP1*, *EMILIN1*, and *PRDM5* are enriched in PG.

**Supplementary Figure S5.**
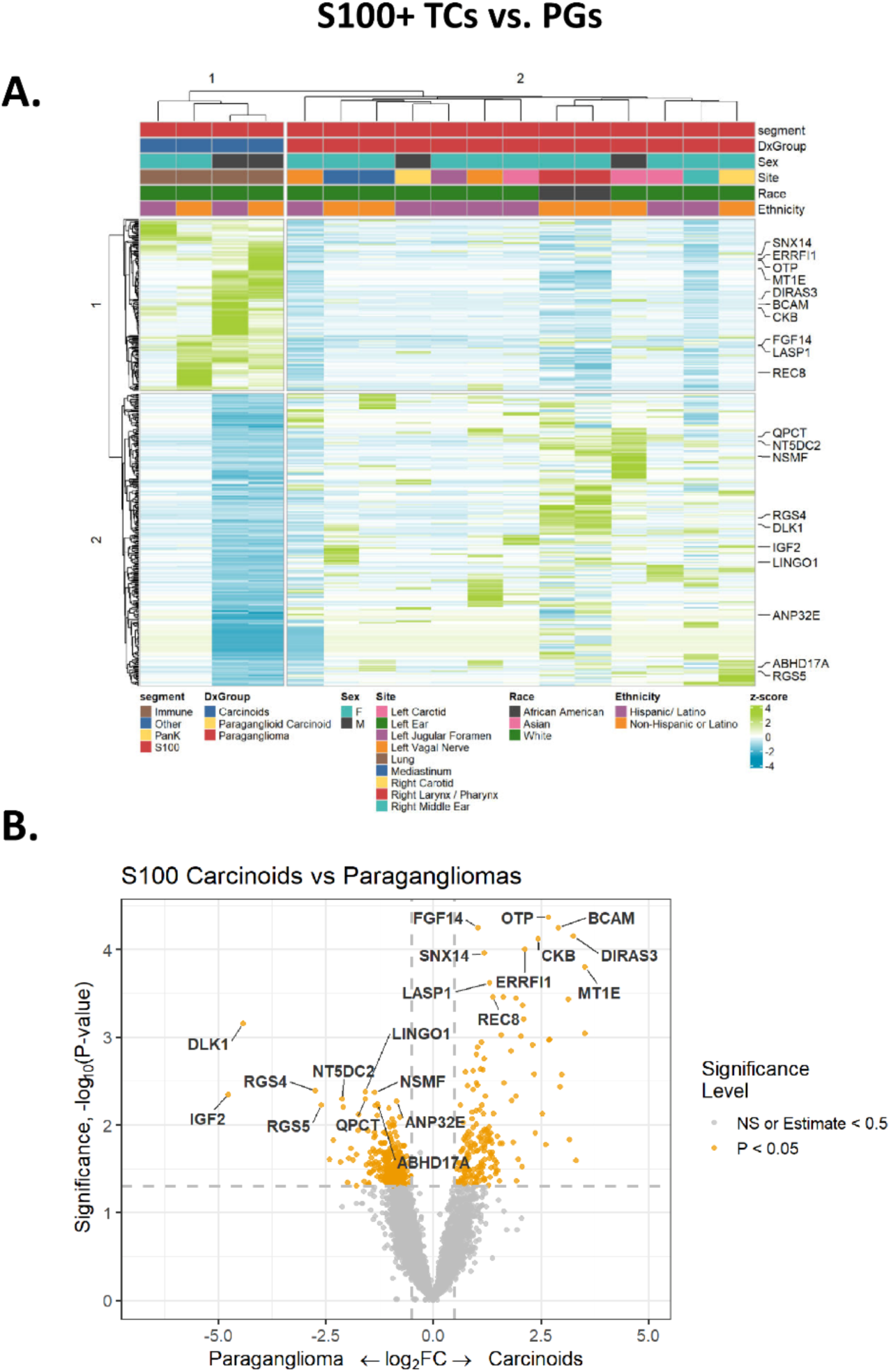
Heat map and volcano plot of the differential gene expression profiles between S100+ TC vs. PG. **(A)** Heatmap displaying the expression of differentially expressed genes between S100-positive PG and TC. Columns correspond to S100+ ROIs, annotated by diagnosis, tissue site, and demographics. **(B)** Volcano plot depicting differential gene expression between S100-positive PG and TC. The x-axis represents the log2-transformed fold change (log2 FC), with positive values indicating higher expression in TC and negative values indicating higher expression in PG. The y-axis represents the –log10-transformed *p*-values, reflecting statistical significance. Nominally significant genes (*p* < 0.05) are in orange and non-significant genes or those with minimal fold change (|log2 FC| < 0.5) are shown in grey. Genes such as *OTP*, *BCAM*, *FGF14*, and *DIRAS3* are upregulated in TC, while *RGS4*, *DLK1*, and *IGF2* are upregulated in PG.

**Supplementary Figure S6.**
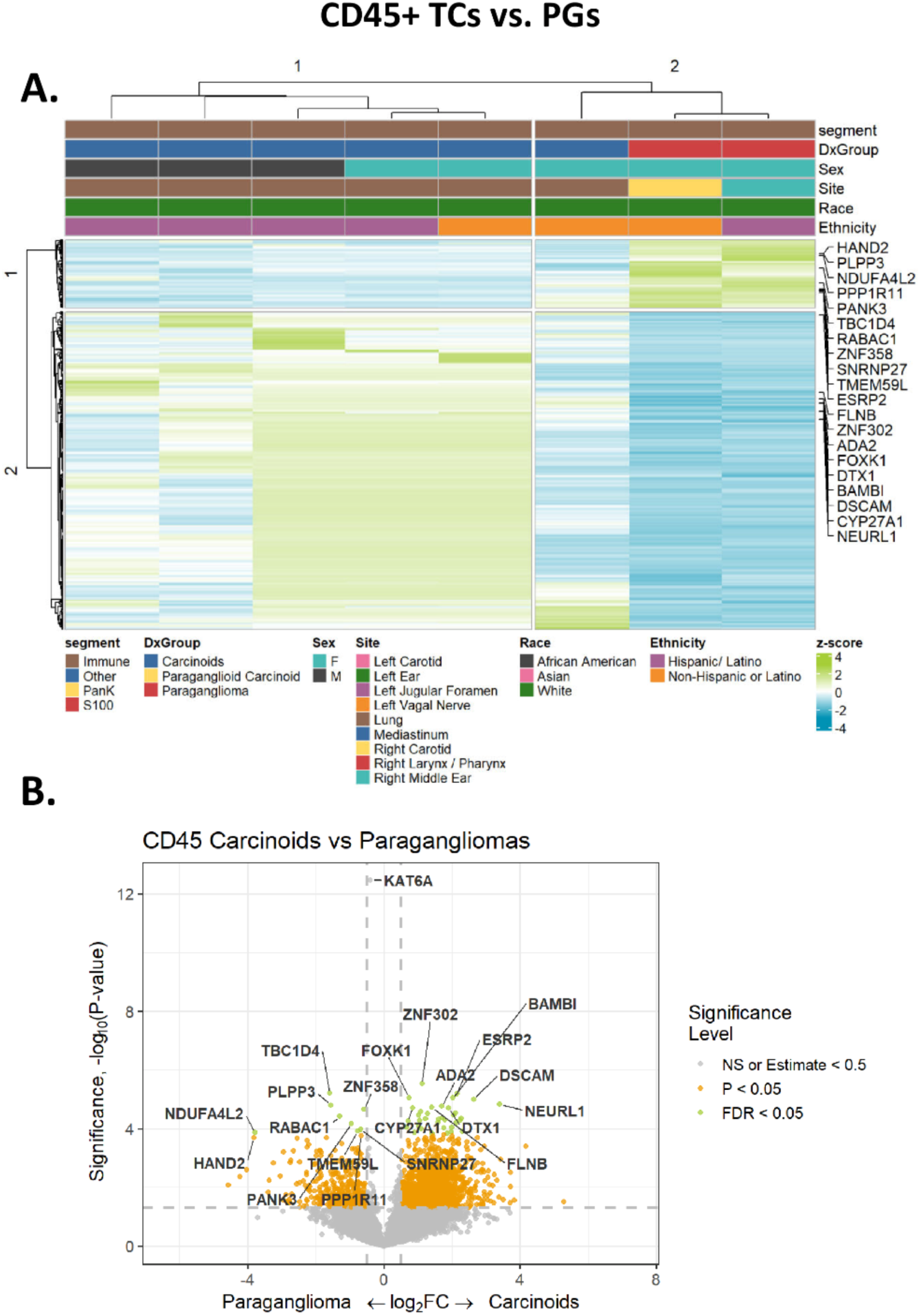
Heat map and volcano plot of the differential gene expression profiles between CD45+ TC vs. PG. **(A)** Heatmap displaying the expression of differentially expressed genes between CD45-positive PG and TC. Columns correspond to CD45+ ROIs, annotated by diagnosis, tissue site, and demographics. **(B)** Volcano plot depicting differential gene expression between CD45-positive PG and TC. The x-axis represents the log2-transformed fold change (log2 FC), with positive values indicating higher expression in TC and negative values indicating higher expression in PG. The y-axis represents the –log10-transformed *p*-values, reflecting statistical significance. Genes with a false discovery rate (FDR) threshold of 0.05 are highlighted in green, nominally significant genes (*p* < 0.05) are in orange, and non-significant genes or those with minimal fold change (|log2 FC| < 0.5) are shown in grey. Notable genes such as *NEURL1* and *DSCAM* are upregulated in TC whereas *RABAC1*, *HAND2*, and *NDUFA4L2* are upregulated in PG.

**Supplementary Figure S7.**
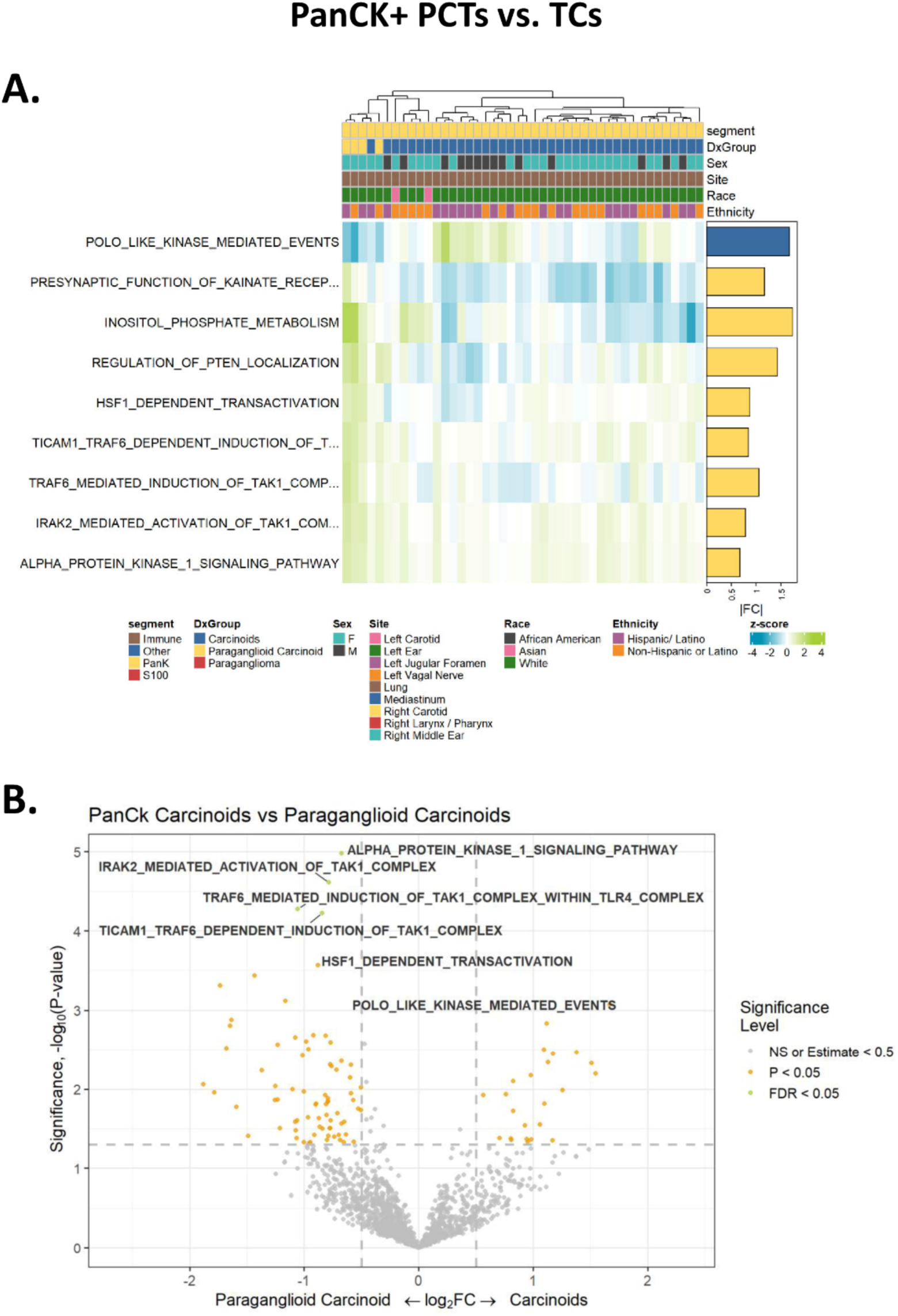
Gene set enrichment analysis comparing PanCK+ PPCT vs. TC. **(A)** Heatmap showing top upregulated and downregulated pathways enriched in PPCT, highlighting activation of inflammatory and membrane signaling cascades. **(B)** Volcano plot of pathway enrichment. Notable enrichment includes *TRAF6*-mediated *TAK1* induction and downregulation of polo-like kinase (PLK) – mediated events.

**Supplementary Figure S8.**
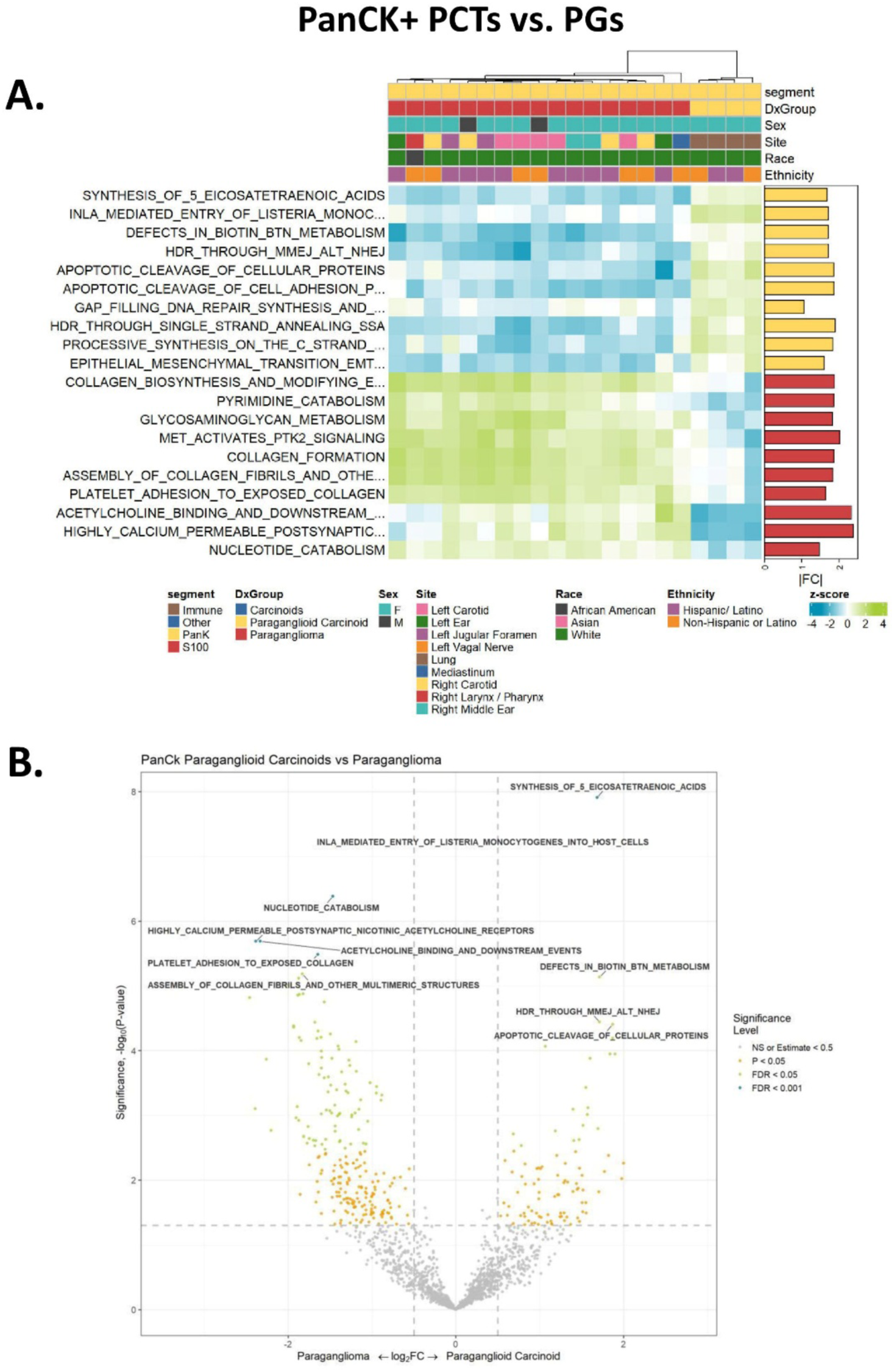
Gene set enrichment analysis comparing PanCK+ PPCT vs. PG. **(A)** Heatmap displaying highly enriched epithelial-associated pathways, including EMT, collagen remodeling, and immune modulation. **(B)** Volcano plot revealing extensive transcriptional divergence, with key upregulation of *NEF*/MHC class I downregulation and membrane excitability pathways.

**Supplementary Figure S9.**
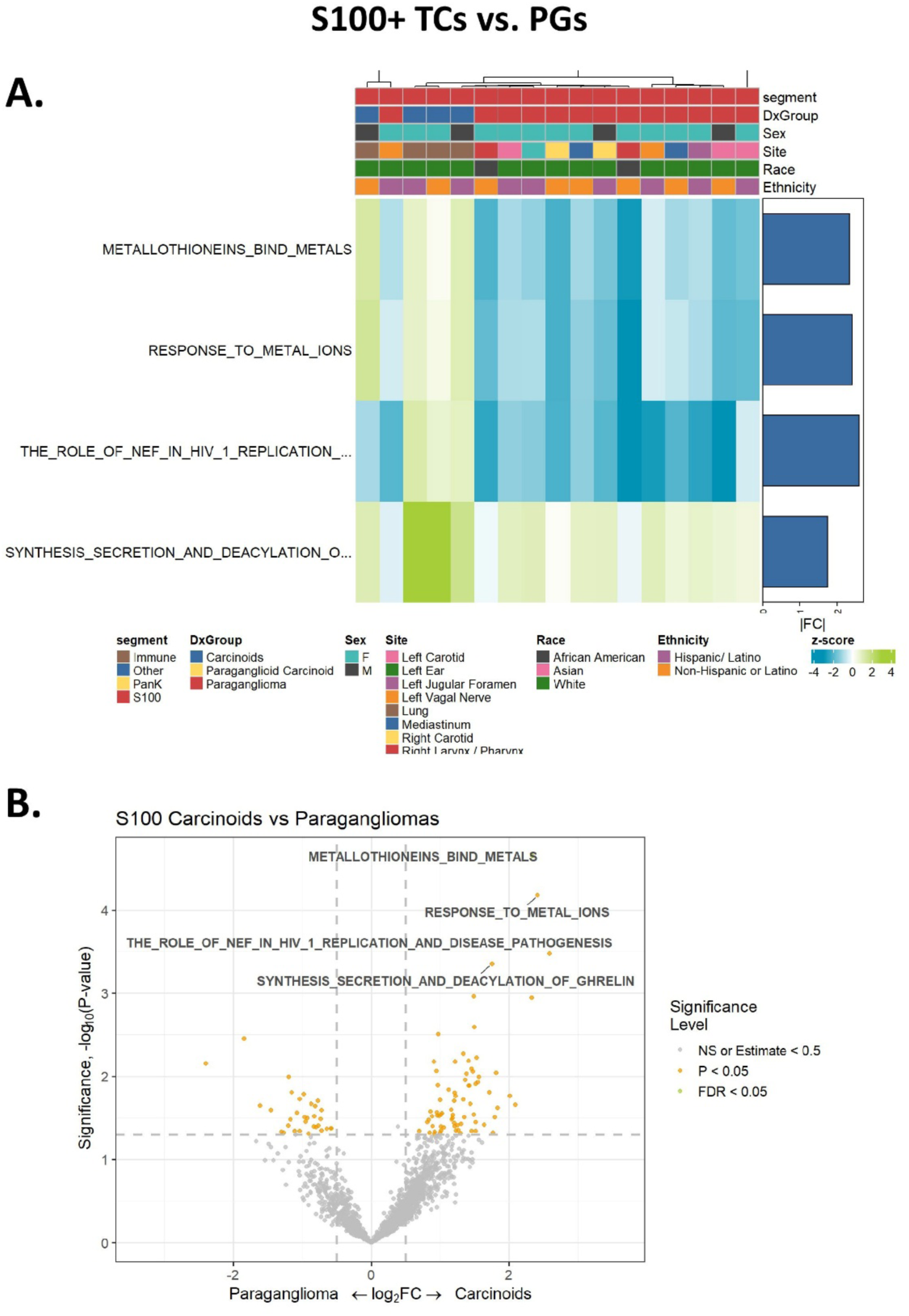
Gene set enrichment analysis comparing S100+ TC vs. PG. **(A)** Heatmap showing limited differentially enriched pathways, primarily related to metal ion metabolism and HIV-1 *NEF* signaling. **(B)** Volcano plot confirms a small number of significant pathway shifts.

**Supplementary Figure S10.**
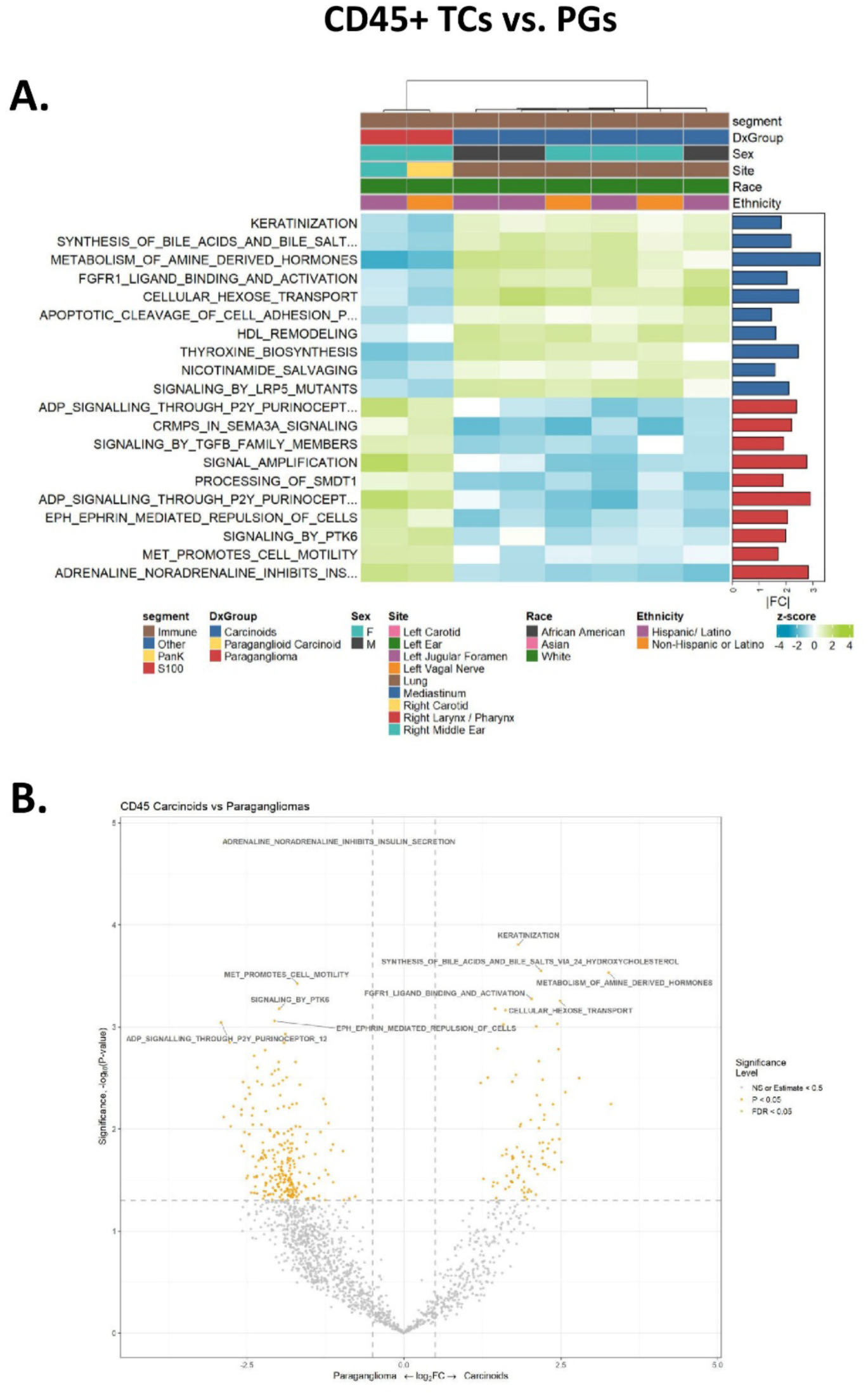
Gene set enrichment analysis comparing CD45+ immune-rich compartments in TC vs. PG. **(A)** Heatmap indicating broad immune-modulatory reprogramming, with altered hormone metabolism, signaling, and cytokine pathways. **(B)** Volcano plot with top enriched and suppressed immune-related pathways, including bile acid, purinergic signaling, and ephrin-mediated repulsion.

## Supplementary Tables

**Supplementary Table S1.** Patient characteristics.

**Supplementary Table S2.** Immunohistochemical and proliferative profiles of TC, PG, and PPCT.

**Supplementary Table S3.** Differentially expressed genes between PanCK+ TC vs. PPCT.

**Supplementary Table S4.** Differentially expressed genes between PanCK+ PPCT vs. PG.

**Supplementary Table S5.** Differentially expressed genes between S100+ TC vs. PG.

**Supplementary Table S6.** Differentially expressed genes between CD45+ TC vs. PG.

**Supplementary Table S7.** Gene set and pathway enrichment analysis between PanCK+ PPCT vs. TC.

**Supplementary Table S8.** Gene set and pathway enrichment analysis between PanCK+ PPCT vs. PG.

**Supplementary Table S9.** Gene set and pathway enrichment analysis between S100+ TC vs. PG.

**Supplementary Table S10.** Gene set and pathway enrichment analysis between CD45+ TC vs. PG.

